# COVID-19 superspreading in cities versus the countryside

**DOI:** 10.1101/2020.09.04.20188359

**Authors:** Andreas Eilersen, Kim Sneppen

## Abstract

So far, the COVID-19 pandemic has been characterised by an initial rapid rise in new cases followed by a peak and a more erratic behaviour that varies between regions. This is not easy to reproduce with traditional SIR models, which predict a more symmetric epidemic. Here, we argue that superspreaders and population heterogeneity are the core factors explaining this discrepancy. We do so through an agent-based lattice model of a disease spreading in a heterogeneous population. We predict that an epidemic driven by superspreaders will spread rapidly in cities, but not in the countryside where the sparse population limits the maximal number of secondary infections. This suggests that mitigation strategies should include restrictions on venues where people meet a large number of strangers. Furthermore, mitigating the epidemic in cities and in the countryside may require different levels of restrictions.

## INTRODUCTION

At its onset the COVID-19 pandemic shocked the world, with the number of new cases and deaths growing more than 20 % per day in the main hotspots [1]. With a growth rate this high, the disease was expected to spread through the population in less than six months without mitigation, and to reach a peak after three months, at which point 30 % of the population would have had the disease [2]. This, however, was not how the initial wave of the epidemic played out [3].

While most of the epidemic undoubtedly was halted due to mitigation efforts, it is striking that even in countries that have implemented a very light lockdown, such as Sweden, the epidemic has peaked long before herd immunity was achieved. Furthermore, even societies that slowly reopened businesses and public life did not experience an immediate explosive resurgence of the epidemic expected given the low levels of immunity and the speed with which the disease spread initially [3].

Here, we will propose an agent-based lattice model of an infectious disease that spreads in a geographically heterogeneous population. The model is a simplified depiction of the dynamics of COVID-19 with its characteristic parameters. We will examine the effect of the heterogeneous infection pattern that is so characteristic of this disease, using a gamma distributed infectiousness with dispersion factor *k* = 0.1 from [4]. Infection heterogeneity is a feature of several epidemic diseases [5], and plays a particularly important role for COVID-19 [4, 6–8]. In individual events, a single person has caused dozens of infections [9]. At the same time, the attack rate within households has been reported to be very low, at less than 20 % [10], despite prolonged close contact. This suggests that the majority of COVID-19 patients infect very little.

In [11] an agent-based model was used to demonstrate that the Achilles heel of an epidemic driven by superspreaders was public social contacts, while the repeated contacts to smaller family and work groups were less dangerous.

Looking at COVID-19 data from the United States in the analysis by [12], it is clear that a rapidly spreading epidemic occurs primarily in densely populated areas. In less densely populated areas, the epidemic onset is delayed, and in rural areas the epidemic never really starts, with most cases appearing to be spillover from the cities. The peak daily per capita mortality in [12] varies by a factor of ten between the most and least densely populated areas in the USA.

The density dependence of COVID epidemics could in principle be explained by the higher chance of meeting infected people in dense areas. However, only 1% of COVID-19 spread was outdoors in China [13], suggesting that only indoor meetings counts. Furthermore, only few infections happens within households [10], suggesting that it is social visits and meetings in confined areas that facilitate infection [14]. We will base our model on these observations.

## MODEL

Our model plays out on a lattice of side *L* with periodic boundary conditions. Each lattice site may either be empty or contain one agent. The agents can be in one of three states, susceptible, infectious, or recovered. Individuals interact with their neighbours with a frequency *f*_*meet*_ which is fixed along with the mean infection probability per meeting to give the desired number of secondary infections, *R*_0_. The distance that agents travel we draw from the distribution 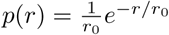 with a mean of *r*_0_ = 10 sites unless stated otherwise. The real probability distribution of travel distances for cars has in Italy been determined to be an exponential function below 20 kilometers, and then a powerlaw with a steeper cutoff around 500 kilometers [15]. The interaction radius *r*_0_ is not by itself a meaningful parameter. Rather, it is the number of neighbours within this radius, given by 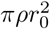 (where *ρ* is the population density) which determines the behaviour of the system, as will be demonstrated below.

COVID-19 is known to be transmitted before the onset of symptoms [16]. For simplicity, rather than using the SEIR modeling framework, we combined the exposed, presymptomatic state and the overlapping infectious period (both estimated at about five days) into one single infectious period of 1*/γ* = 10 days [17, 18]. Infected agents randomly leave the infectious state with a probability of *γ* per day, meaning that the duration of the disease is exponentially distributed.

When an infectious agent *i* interacts with a susceptible agent, the susceptible agent will become infected with a probability *p*_*i*_ that is specific to the infecting agent. We draw these probabilities from a gamma distribution with dispersion factor *k* = 0.1 [4, 5], within the range observed for COVID-19 [4, 6, 8]. The distribution is normalised to give an average reproduction number *R*_0_ of 3 at a population density of 1.

The geographical heterogeneity of the population is modeled by placing a square “city” of side *L/*5 on a lattice with periodic boundary conditions and lattice size *L*. The city has the population density 1, i.e. all sites are occupied, and the city population is thus *L*^2^/25. The city is surrounded by “countryside” with a population density *ρ* and total population of 24*ρL*^2^/25.

Importantly we assume that the rate of interactions per agent is kept the same in both city and countryside, meaning that we assume that people are equally social. If an agent in the countryside attempts to interact with an empty site, the attempt is counted as failed, and new attempts are made until the number of contacts is the same as in the city, where all sites are occupied.

Thus, people in the countryside interact with a smaller set of people while still spending the same amount of time on social activities. In a wider perspective this proposes that density dependence of disease spreading is more due to difference in diversity of contacts than due to differences in time spent around other people. Thereby our model assumes an infection rate that depends on density, but not in a simple linear fashion as sometimes assumed [19].

We seed the disease within the city. Even if we were to seed it randomly, the city would usually be hit early on provided that the epidemic catches on.

## RESULTS

In fig. 2 we consider a disease with heterogeneous infection rates and study how simulated epidemic trajectories depend on population density. One sees that the dynamics resembles that of a SIR model at *ρ* values much lower and much higher than the critical density. In the low regime, the epidemic only spreads in the city which is relatively well-mixed, whereas in the high regime the countryside begins to resemble the city more and more. In both extremes, the fraction of infected individuals rise and fall symmetrically. The figure illustrates how the epidemic is “stretched out” in the intermediate density range. The epidemic has the longest lifetime when *ρ* is just above the percolation threshhold *ρ*_*crit*_, as the disease still spreads in the countryside, but is nonetheless slowed down by te lower population density.

**FIG. 1.**
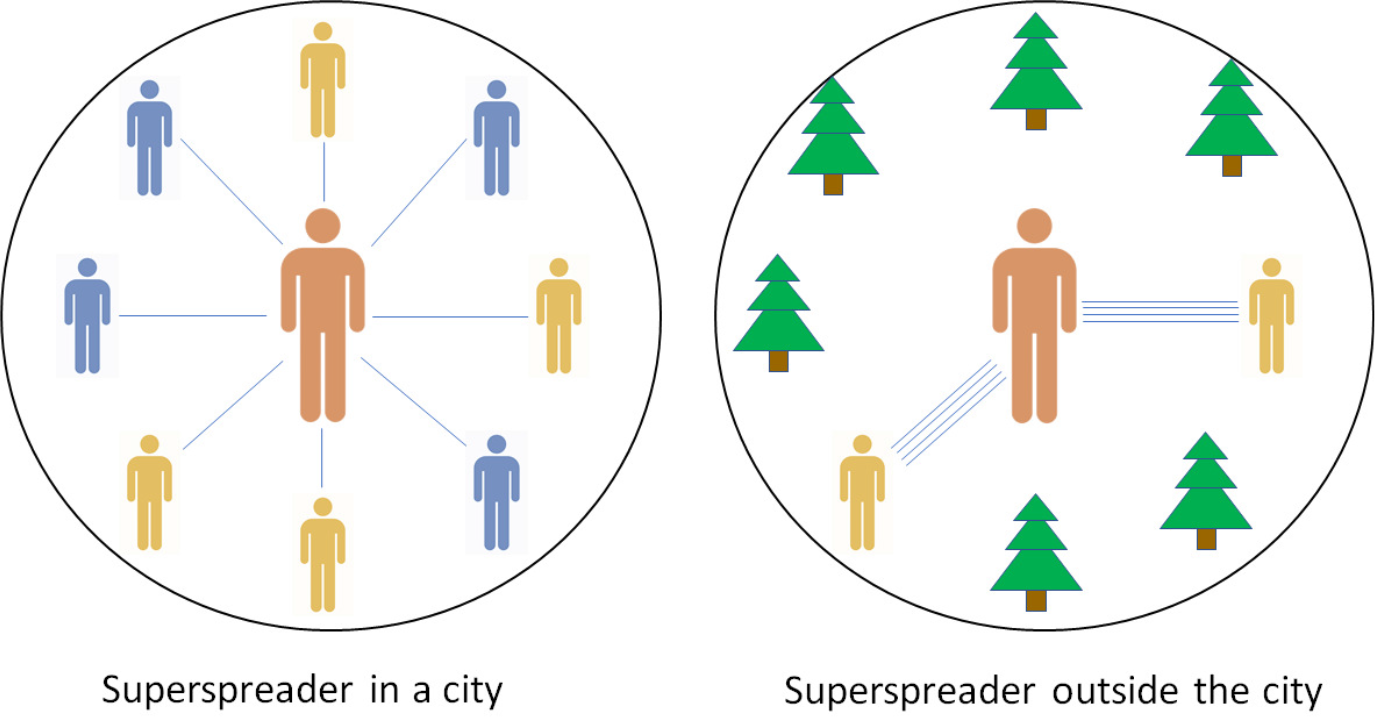
Model: A superspreader in a city interacts a little with a lot of people and will infect some fraction of them. On the other hand, a superspreader outside the city will interact a lot with each of a smaller set of people. The superspreader then infects practically all of them, but there is a lower cap on the number of secondary infections.

**FIG. 2.**
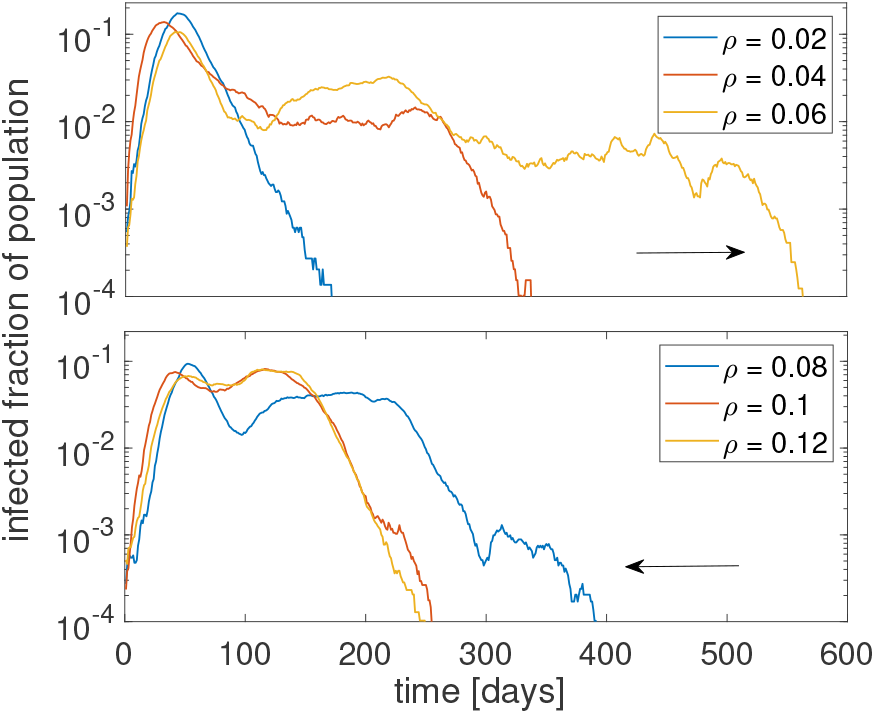
Epidemic trajectories: Infected fraction of the population over time changes with the countryside population density *ρ*. In the low-*ρ* regime, increasing the population density stretches the curve, as the epidemic spreads further from the city. When *ρ* is above ∼0.06, the epidemic again approaches the behaviour of a SIR model, as the epidemic now spreads unhindered across the whole system. Around *ρ*_*crit*_, there is a large variation in the duration of the epidemic. The parameters used are *γ* = 0.1, *r*_0_ = 10, *f*_*meet*_ = 10 and dispersion parameter *k* = 0.1.

In fig. 3 (a, b) we measure the attack rate of the epidemic in a homogeneously distributed population in order to find the percolation threshhold, below which the epidemic will stop propagating. Panel (a) identifies *ρ*_*crit*_ ≈0.01 for a homogeneously spreading disease. In contrast, a disease with an overdispersion of *k* = 0.1 has a much higher critical density *ρ*_*crit*_ ≈0.04, as seen in (b). The same figure demonstrates that it is not the density alone that determines the ability of the disease to percolate, but rather the number of neighbours, proportional to 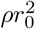. As expected, the disease with homogeneous infectivity can spread already when each person has one neighbour on average, whereas the overdispersed disease requires almost four neighbours to percolate. The overdispersed simulation and the simulation with homogeneous infection were done with same average disease transmission rate and the factor ∼4 difference in critical density comes about because a disease with *k* = 0.1 has 10 % of the infected being responsible for 80 % of the infections. Thus most people do not transmit the disease, and it is therefore the density of the few people who do spread the disease that sets the critical threshhold.

**FIG. 3.**
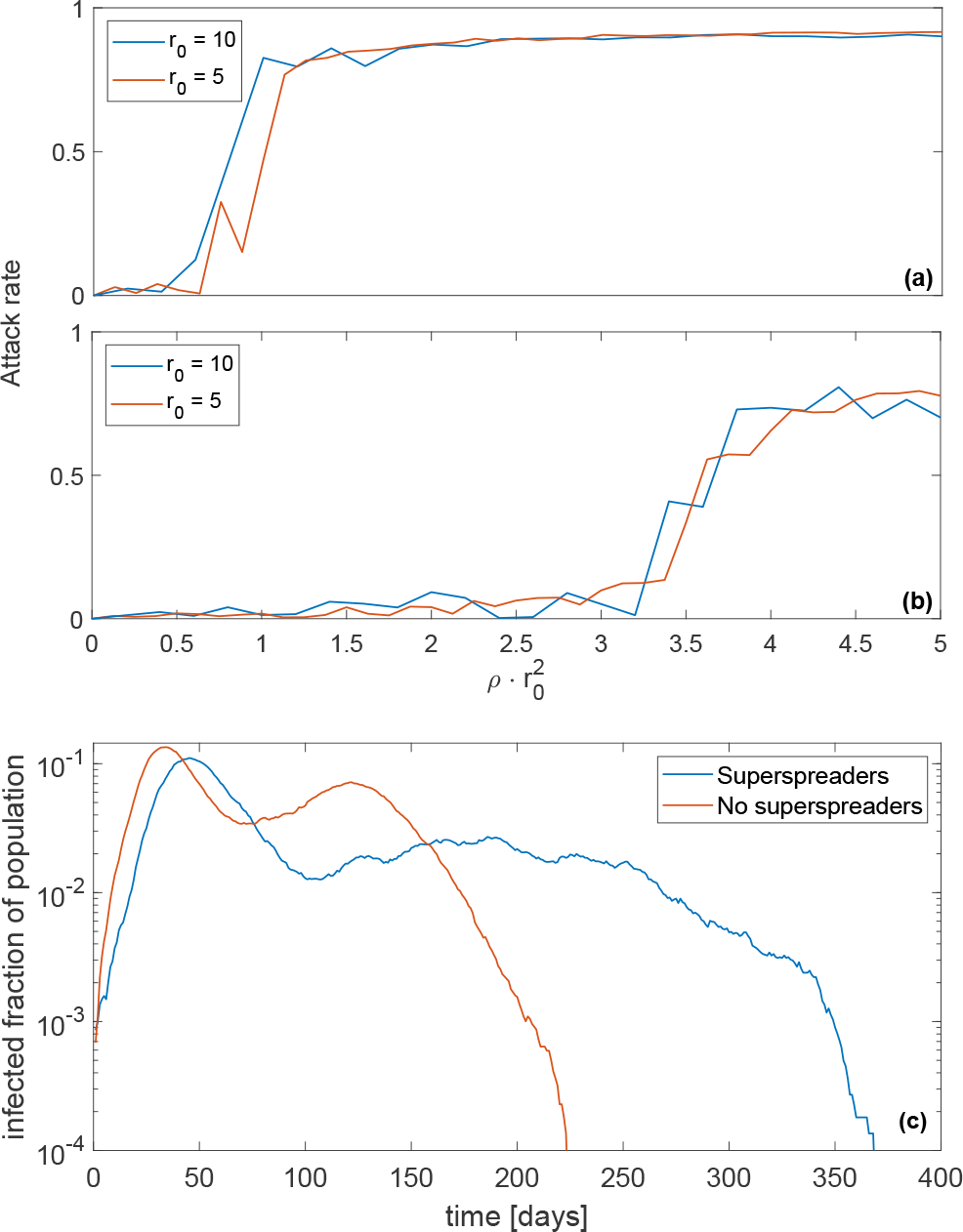
Comparison of models without and with superspreaders. Panel (a) shows the attack rate as a function of the number of neighbours within the radius of interaction 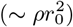 in a population where everyone infects with the same rate. (b) shows attack rate with heterogeneous infection rates, using a gamma distribution with dispersion factor *k* = 0.1. The two overlaid curves demonstrate that the parameter *r*_0_ does not affect the physics of the system, and what really determines the ability of the disease to percolate is the number of neighbours, proportional to 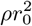. (c) Epidemic trajectory when superspreaders dominate (blue) and when infectiousness is evenly distributed (red) for equal countryside population density and radius of interaction 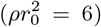. When superspreaders are the main drivers of the epidemic, it is strongly impeded once the city has reached herd immunity. When everyone infects equally, the epidemic simply spreads radially out from the city, leading to a “second wave” in the countryside. Parameters are as in fig. 2.

Since our model analysis centers on superspreaders as a main driver of the epidemic, fig. 3(c) compares epidemics with and without superspreaders. It can be seen that with no superspreaders, the epidemic will spread un-hindered in the countryside, albeit more slowly since the countryside is geographically larger than in the city.

This leads to a graph similar to two superimposed SIR-like models. If superspreaders are present, however, the epidemic may spread both slowly and erratically in the countryside and continue long after herd immunity is achieved in the city.

The dependence of the percolation threshhold on the dispersion parameter *k* is further investigated in fig. 4, which shows the attack rate as a function of both the number of neighbours and *k*. The figure shows that the percolation threshhold increases drastically at low *k*, meaning that a superspreader-driven epidemic requires more social contacts per person in order to spread. Once the epidemic becomes sufficiently overdispersed (*k <* 0.05), it is no longer viable.

**FIG. 4.**
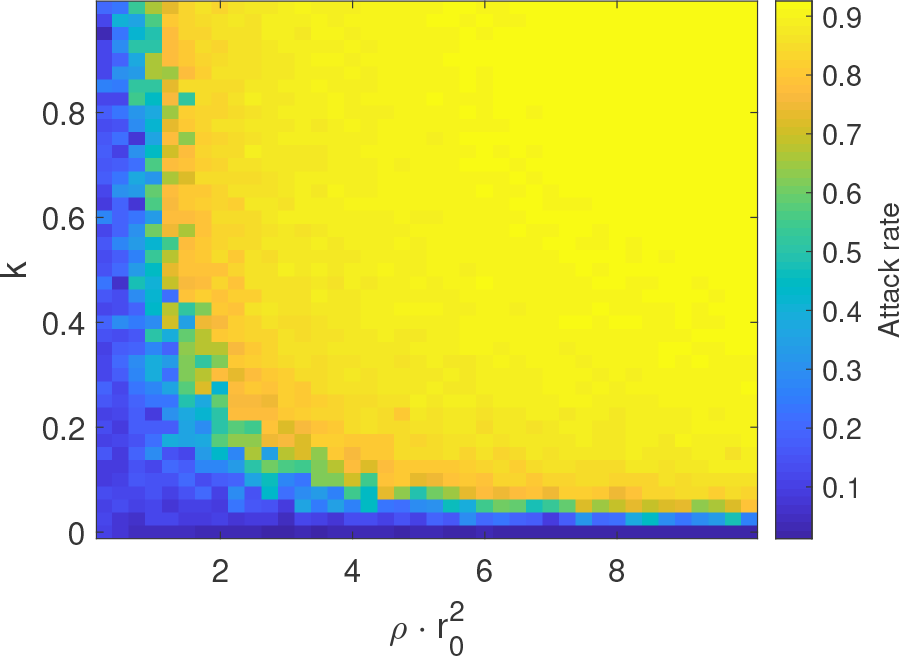
Dependence of attack rate on density and *k*. Since the variable determining percolation is not the absolute density, but the number of neighbours, we plot 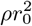 on the x-axis rather than *ρ*. It is seen that the disease percolates much more easily at a higher *k*, which implies a more homogeneous infectivity. The more overdispersed the disease (corresponding to lower *k*), the more neighbours are required for percolation.

In fig. 5, we try to replicate the data compiled by [12] and see that local disease incidence in a model with heterogeneous infectivity is indeed much more population density dependent than a model assuming homogeneous infectivity. This fits well with the cited data, which suggest a strong dependence of COVID-19 incidence on population density. It has already been known for years that the spread of epidemics is population density dependent [19]. Here, we show that this dependence is enhanced by heterogeneous infectivity. Importantly, as opposed to traditional disease models, we assume that everyone is equally social, but that the set of available contacts is smaller in sparsely populated regions. The significance of this will be discussed further below.

**FIG. 5.**
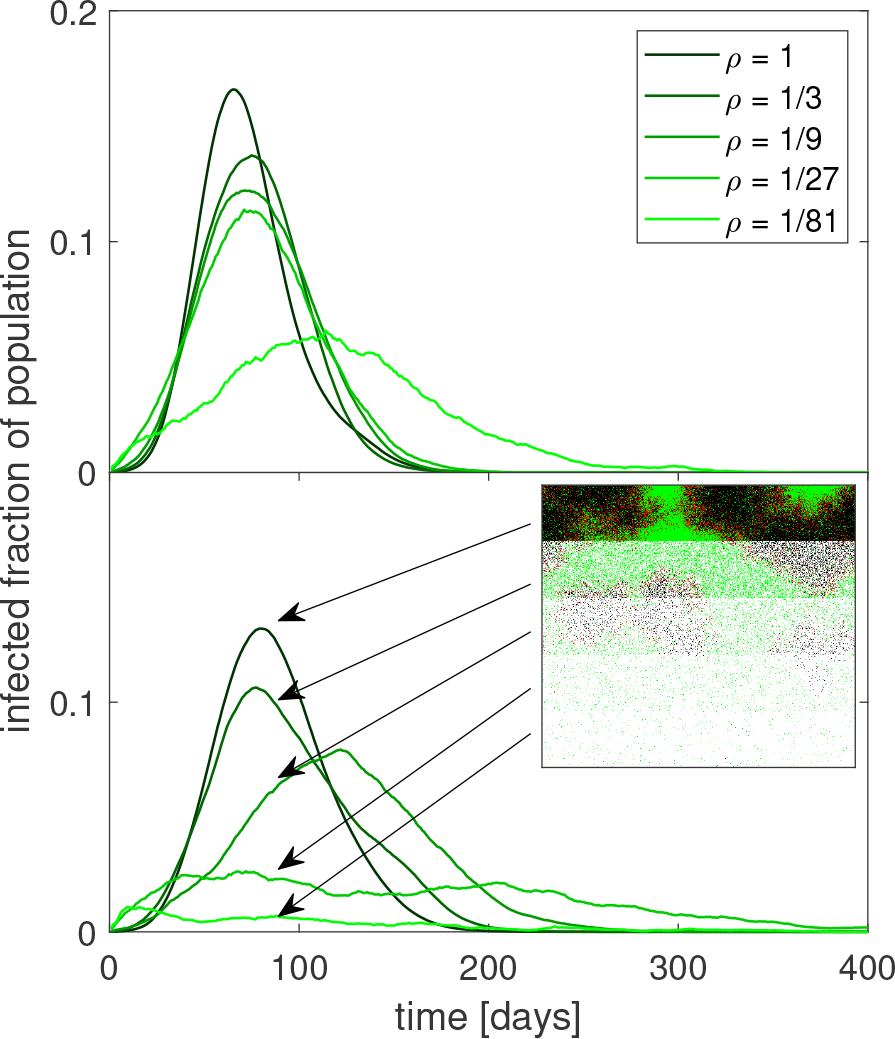
Epidemic trajectory in regions of varying density. When infectivity is homogeneous (top), the epidemic is a lot less sensitive to a lower population density than when the epidemic is driven by superspreaders (bottom). Here, the epidemic is nearly absent in the low-density regions, and appears to be driven by spillover. Inset illustrates the layout of the lattice.

**FIG. 6.**
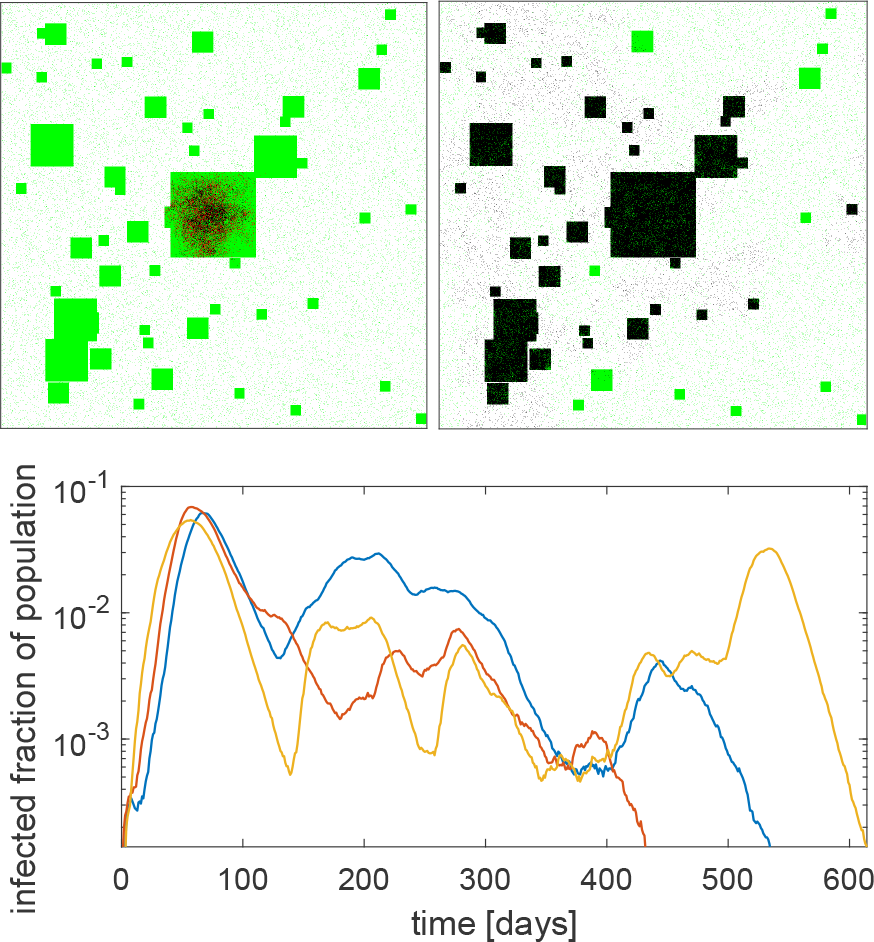
Simulation with multiple cities: Infected individuals are shown in red, susceptibles are green, empty sites are white and recovered are black. City size distribution mimics Zipf’s law [23], such that there is one city of with 40000 inhabitants, 4 with 10000, and so forth. The graphs show the fraction of the population currently infected as a function of time for three example runs of the simulation. Here, *ρ* = 0.03 and the other parameters are the same as the above figures.

The delayed onset and erratic behaviour of the countryside epidemic obviously depends on the density and other characteristics of the countryside and the assumed travelling pattern of individuals. Also, real countryside contains a diverse pattern of smaller and large settlements. Therefore we considered a system with several cities distributed on a lattice with side length *L* = 1000. The system mimics the observed city size distribution which is fairly close to the Zipf law [20]. The random distribution of smaller cities is not entirely naturalistic, since evidence suggests that real cities are organised in a fractal pattern [21, 22]. A figure using a city distribution closer to a fractal can be found in the supplement. For illustrative purposes we chose a density in the countryside that is close to but below the percolation threshhold for the disease (*ρ* = 0.03). With this below-critical spreading we observe cities that are spared and cities that are nearly completely infected. In reality assuming near critical spreading in the countryside would not be not necessary for global spread, since people occasionally travel long distances [15], facilitating rare direct transmissions between distant cities.

## DISCUSSION

Based on the above, we suggest that the lopsided appearance of COVID-19 epidemic curves can be explained by heterogeneous infection ability combined with a geographically heterogeneous population. However, our model makes a number of assumptions and breaks from reality, whose importance must be discussed.

First and foremost, the distribution of full and empty lattice sites does not represent the geographical distribution of people as such. Rather, it represents the density of contacts. The density ratio between people in the countryside compared to an inner city is much lower than used in our model. On the other hand, persons living outside the cities may be more mobile than city-dwellers. The density contrast of agents on our lattice represents the combined effect of these factors.

A further complication is the distribution of disease duration and incubation periods. We here assume a simple exponential distribution of infectious period duration, whereas the real mechanics of COVID-19 includes presymptomatic transmission and a broad gamma distribution of incubation periods [17]. A different infectious period distribution might complicate our findings. Therefore, we examine the effect of a gamma distributed infectious period duration in the supplement. We find that, while changing this distribution has an effect on the percolation threshhold, it does not change our fundamental conclusions.

It has long been discussed how disease transmission rate depends on population density [19, 24, 25]. We here present a new way of looking at this problem. A classical SIR model with pseudo-mass action transmission would assume a simple linear dependence of *R*_0_ on population density, implicitly assuming that people become more social by living in a densely populated area.

Our model makes a different assumption: People will be equally social regardless of population density, but when population density is lower, the groups that they spend time with will be less diverse. The number of possible unique contacts for each individual declines linearly with population density, but people in sparsely populated areas are likely to have multiple encounters with the same persons, as their communities are smaller.

If, however, the epidemic is dominated by superspreaders who only need one or a few encounters to transmit the disease, the duration of each encounter becomes less important as even a rather brief contact to a superspreader is likely to lead to infection. Instead, what limits the action of superspreaders is their number of unique contacts. Superspreaders that interact with only a small, tight-knit group will inherently be highly limited in how many secondary infections they can generate.

If superspreading makes a disease vulnerable to variations in population density, we should conversely expect to see that diseases with a homogeneous infectivity, i.e. a dispersion factor close to or above 1, exhibit little variation with population density. One example of a disease with a homogeneous infectivity is influenza, with an estimated dispersion factor of *k* = 0.94 for the 1918 pandemic flu [26]. When examining the incidence of seasonal influenza, which we assume to have a similar dispersion factor, [27] find no consistent variation with population density. This is a point in favour of the link between superspreading and population density dependence.

Finally, the effect of lockdowns and changes of social behaviour is important. A previous paper [11] suggests that even moderate mitigation may limit the action of superspreaders by reducing the maximal number of people any person can be in contact with. If this hypothesis is true, bans on gatherings and a reduction in public social life would lead to early peaks in the number of new cases. Our study compounds this finding, and suggests that a change in behaviour is not strictly necessary to cause an epidemic peak well before herd immunity has been achieved. Mitigation strategies that primarily target cities may well be sufficiently effective in bringing down the epidemic. However, large events like funerals, weddings, or festivals are not included in our model and will of course facilitate spreading in any location.

Despite some caveats, our model reproduces the main aspects of geographical heterogeneity and suggests a new view on density dependence of epidemic dynamics. An epidemic with a large heterogeneity in infection rates is predicted to be most intense in large cities while it slowly tapers off in the countryside. This is consistent with what we see in data from the COVID-19 pandemic [12, 28]. Our results thus favour the hypothesis that the COVID-19 pandemic is driven by superspreaders, and that the observed quick exponential growth phase, early peak, and slow recovery phase are consequences of combining heterogeneity of infectivity with a heterogeneous population density.

## Data Availability

An implementation of the model described in this article can be found on figshare under the DOI
https://doi.org/10.6084/m9.figshare.12919709.v1

## ACKNOWLEDGMENTS

We thank Lone Simonsen, Viggo Andreasen, and Bjarke Frost Nielsen for enlightening discussions. Our research has received funding from the European Research Council (ERC) under the European Union’s Horizon 2020 research and innovation programme under grant agreement No. [740704].

## supplement

### Different distribution of infectious periods

In order to take into account that the duration of illness caused by COVID-19 does not actually follow a neat exponential distribution, we have here considered the case where the duration of infection is gamma distributed as well, though with a dispersion factor greater than one (*k* ≈4). We repeat the central figures 3 and 5 from the main article in order to see if a different distribution of infectious period durations changes our results. The gamma distribution should give fewer people with a very short disease duration, though both distributions have a relatively long tail. The mean duration of infectiousness is the same as in the main article (10 days). An important complication, that individuals with a long period of illness are likely to be hospitalised or otherwise isolated for part of it and thus less infectious, has not been accounted for here.

We find that, while the different distribution of infectious periods does lower the percolation thresh-hold slightly and also diminishes the population density dependence of the epidemic, it does not fundamentally change our conclusions.

**Figure 1:**
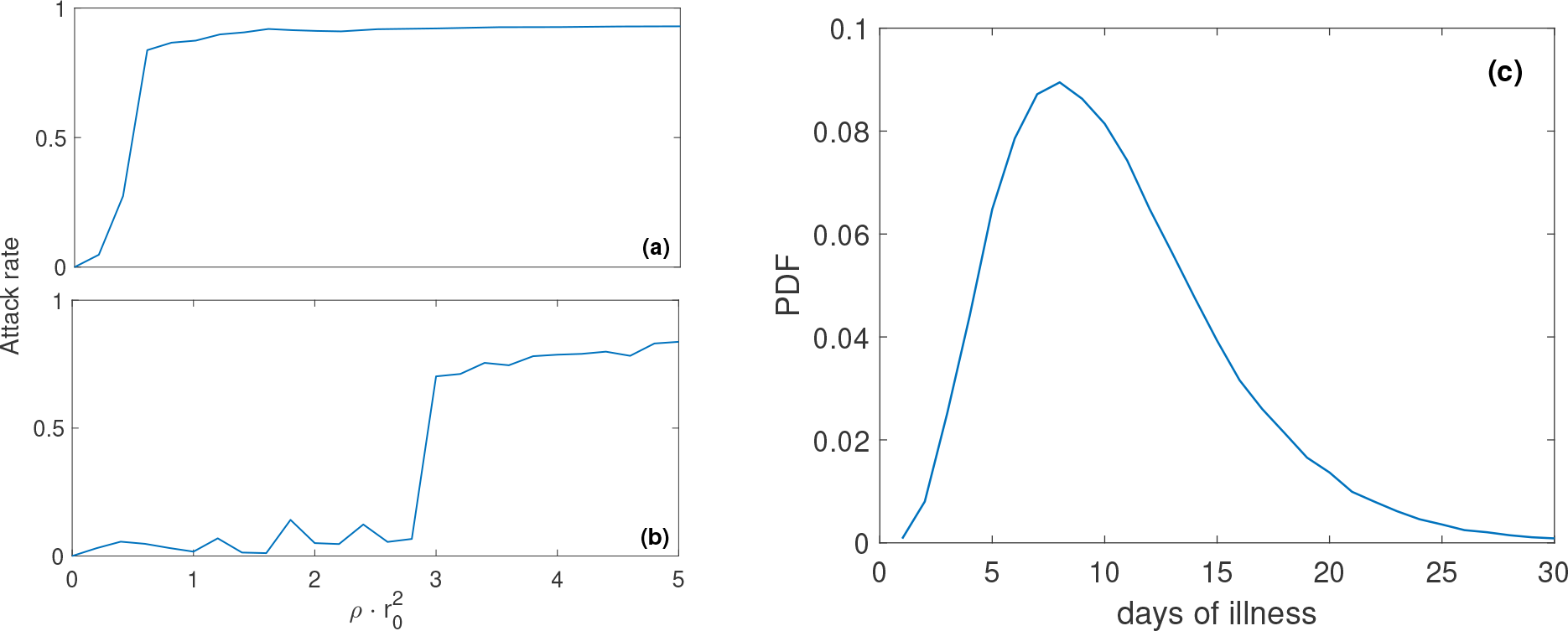
(a) and (b) show attack rates as a function of 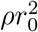, a measure of the number of neighbours within a radius of *r*_0_. (a) shows the case with no infectivity overdispersion (*k* = 1) and (b) shows the case where infectivity is overdispersed with *k* = 0.1. We see that the system behaves similar to the main model, but with a slightly lower percolation threshhold in both cases. (c) shows the probability distribution function for disease duration used here.

**Figure 2:**
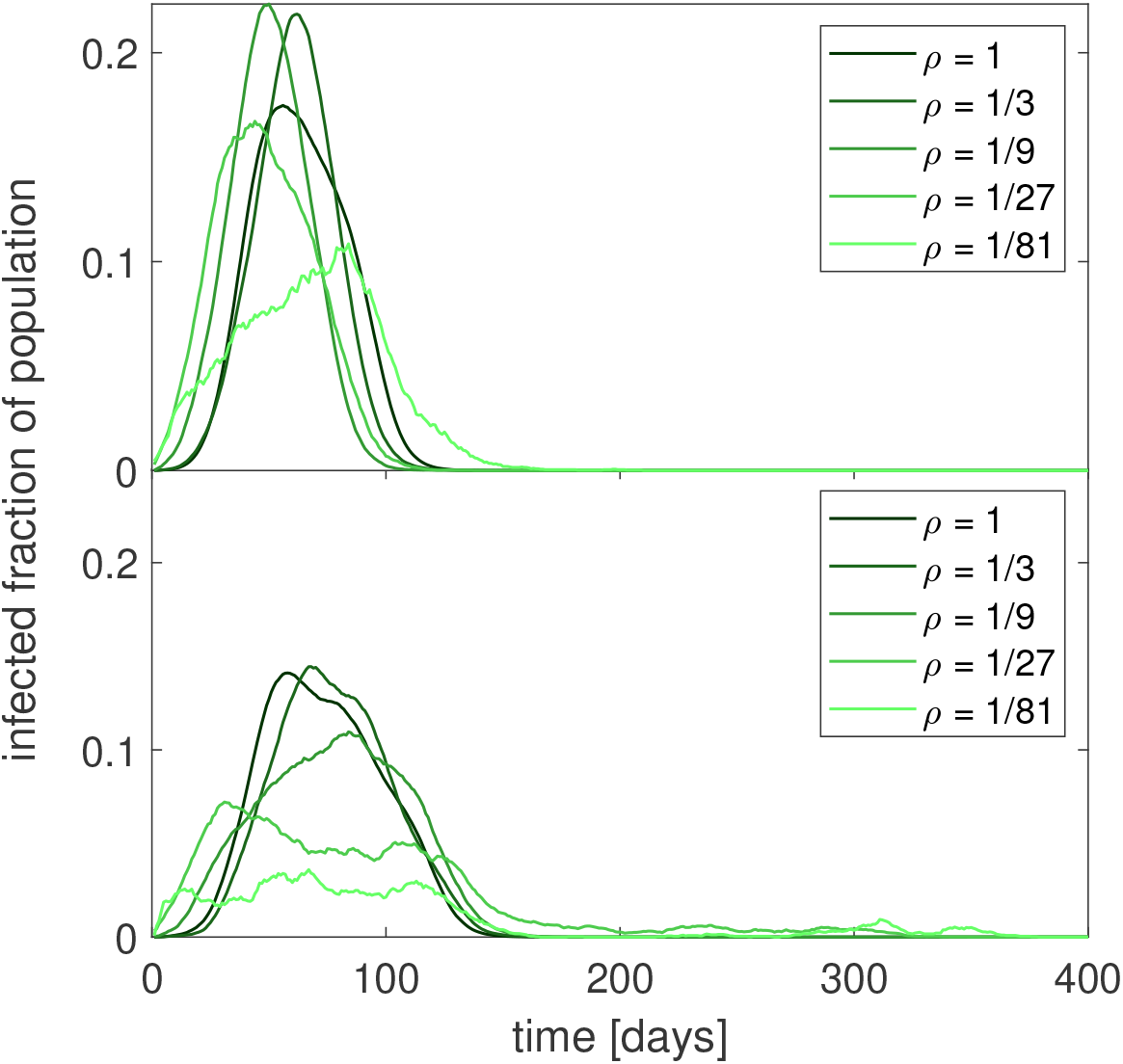
A comparison of the effect of population density on an epidemic that is not overdispersed (top) and an overdispersed epidemic (bottom). As above, the duration of illness is gamma distributed. The effect of density is somewhat reduced in the overdispersed case, but still noticeably greater than the non-overdispersed case. The smaller number of people with very short disease durations using this distribution seemingly also helps the epidemic spread more effectively.

### Fractal city distribution

The random distribution of cities used in fig. 6 of the main article is not entirely realistic. Rather, there is evidence suggesting that real geographical distribution of cities follows a fractal pattern [1,2]. We therefore repeat the figure here with a (partially) fractal distribution of cities. The results of this simulation are fairly similar to the random city distribution.

**Figure 3:**
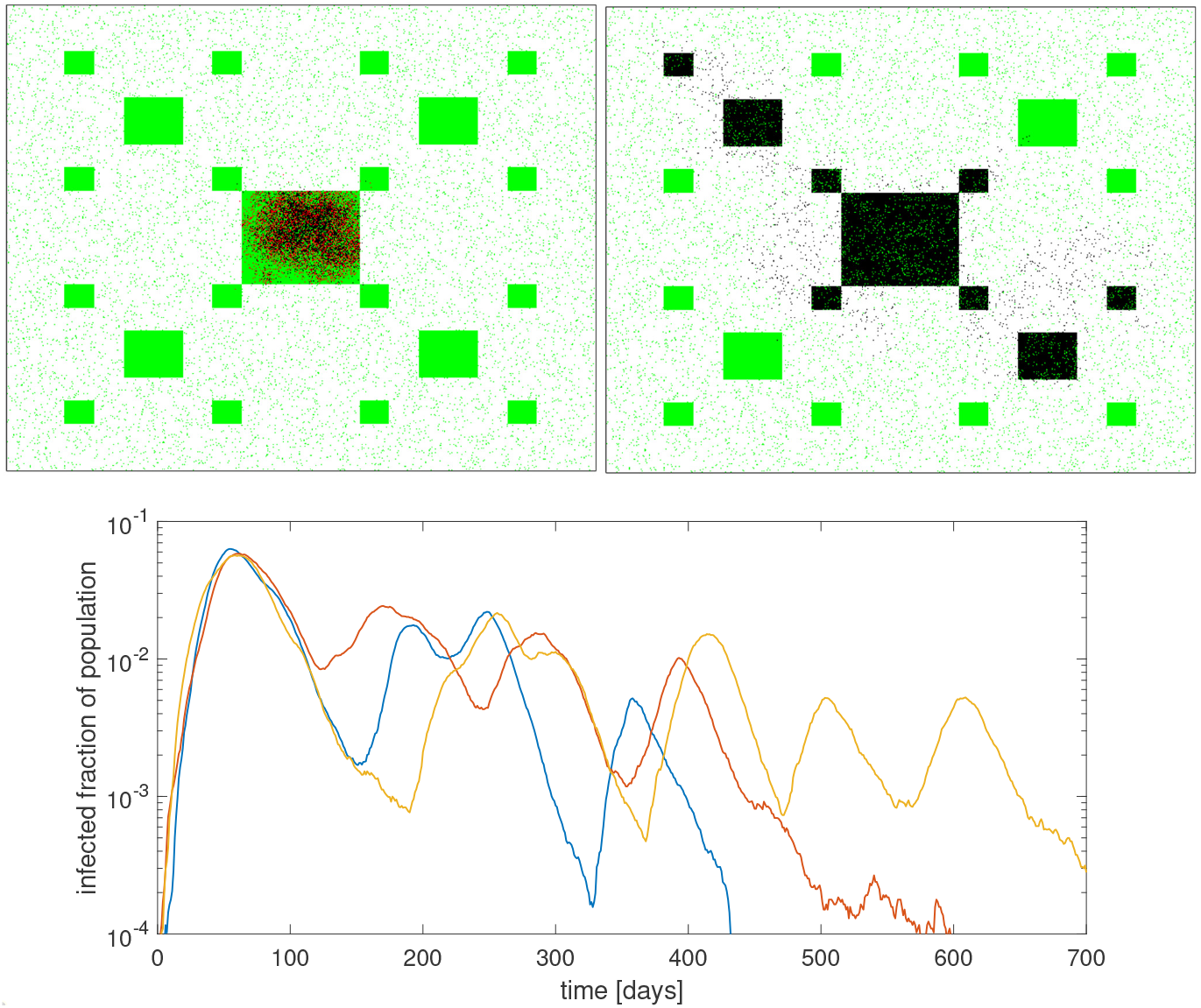
A simulation where, instead of a random distribution, the cities are laid out in a fractal pattern. We see that this makes little difference from the random distribution shown in the main article. The lower panel shows epidemic trajectories from three test runs.

